# Adverse Events After LP.8.1-Containing COVID-19 mRNA Vaccines

**DOI:** 10.64898/2026.01.25.26344612

**Authors:** Anders Hviid, Emilia Myrup Thiesson, Niklas Worm Andersson

## Abstract

**Background:** The LP.8.1-containing COVID-19 mRNA vaccines were recommended for the 2025 seasonal vaccination campaigns in Europe and the United States. Safety data on these vaccines are limited.

**Methods:** We conducted a nationwide register-based cohort study in Denmark including all adults aged 65 years and older or at high risk of severe COVID-19 who had received previous COVID-19 vaccine doses. The study period was July 1, 2025, to December 3, 2025. We estimated incidence rate ratios using Poisson regression comparing rates of 30 adverse events within 28 days following LP.8.1-containing vaccination with reference period rates, adjusted for age, sex, region of residence, high risk of severe COVID-19, calendar time, and comorbidities. Self-controlled case series analysis was conducted as a complementary approach.

**Results:** Among 1,565,697 individuals (mean age 69.5 years; 53.8% female), 958,633 received an LP.8.1-containing vaccine. Receipt of an LP.8.1-containing vaccine was not associated with a statistically significant increased rate of any of the 30 adverse events within 28 days after vaccination. The incidence rate ratio was 0.95 (95% CI, 0.86-1.06) for ischemic cardiac event, 0.83 (95% CI, 0.76-0.92) for cerebrovascular event, and 0.32 (95% CI, 0.04-2.50) for myocarditis. Results from the self-controlled case series analysis were similar.

**Conclusions:** In a nationwide cohort of more than 1.5 million adults, no increased risk of 30 adverse events was observed following vaccination with LP.8.1-containing COVID-19 mRNA vaccines.

## Introduction

The LP.8.1-containing COVID-19 mRNA vaccines were recommended for the 2025 seasonal vaccination campaigns in Europe and the United States.^1,2^ In Denmark, the LP.8.1-containing vaccines were recommended to individuals aged 65 years and older or those at high risk of severe COVID-19, beginning October 1, 2025. Safety data on LP.8.1-containing vaccines are limited. We investigated the associations between LP.8.1-containing vaccines and risks of 30 adverse events.

## Methods

A study cohort of all adults in Denmark aged 65 years and older or belonging to a high-risk group who had received previous COVID-19 vaccine doses, was constructed by linking nationwide health and demography registers at the individual level. The study period was July 1, 2025, to December 3, 2025, and vaccination status was classified in a time-varying manner (eTable). The 30 adverse events were selected from prioritized lists of adverse events of special interest to COVID-19 vaccines and previous research.^3,4^ Each outcome was studied separately and identified as any first hospital contact where an outcome diagnosis was recorded.

Incidence rate ratios were estimated using Poisson regression, comparing outcome rates within the risk period of 28 days following LP.8.1-containing vaccine administration with reference period rates from day 43 after a vaccine dose (both LP.8.1-containing and previous vaccine types) and onward (eFigure). Models were adjusted for age, sex, region of residence, high risk of severe COVID-19, calendar time, and number of comorbidities. This was complemented by a self-controlled case series (SCCS) analysis comparing the predefined risk and reference periods as in the Poisson regression within the same individuals.^5^ A 95% CI that did not cross 1 was defined as statistically significant. Analyses were performed as surveillance activities of the governmental institution Statens Serum Institut, Copenhagen, Denmark. According to Danish law, national surveillance activities conducted by Statens Serum Institut do not require approval from an ethics committee.

## Results

Among 1,565,697 individuals vaccinated in previous seasons (mean [SD] age, 69.5 [13.8] years; 53.8% female), 958,633 received an LP.8.1-containing COVID-19 mRNA vaccine during follow-up (Table). In the Poisson analysis, receipt of an LP.8.1-containing vaccine was not associated with a statistically significant increased rate of hospital contacts for any of the 30 adverse events within 28 days after vaccination compared with reference period rates (Figure). The incidence rate ratio was 0.95 (95% CI, 0.86-1.06) for ischemic cardiac event, 0.83 (95% CI, 0.76-0.92) for cerebrovascular event, and 0.32 (95% CI, 0.04-2.50) for myocarditis. Results from the SCCS analysis were similar, with no statistically significant increased risks observed. Some outcomes were very rare during follow-up resulting in low statistical precision. However, for the majority of adverse events examined, the upper bound of the 95% CI was inconsistent with moderate to large increases in relative risk.

**Table:** Cohort Characteristics in a Study of LP.8.1-Containing Vaccines^a^.

**Table: Cohort Characteristics in a Study of LP.8.1-Containing Vaccines<sup>a</sup>**
| **Characteristic** | **Entire study cohort<sup>b</sup>** | **Vaccinated with LP.8.1-containing vaccine** |
| --- | --- | --- |
| Total vaccinated | 1,565,697 | 958,633 |
| Age, mean (SD), yrs | 69.5 (13.8) | 74.3 (9.3) |
| **Sex** |  |  |
| Female | 842,291 (53.8) | 515,483 (53.8) |
| Male | 723,406 (46.2) | 443,150 (46.2) |
| **Region of residency in Denmark<sup>c</sup>** |  |  |
| Capital | 428,321 (27.4) | 261,227 (27.2) |
| Central | 353,887 (22.6) | 221,468 (23.1) |
| Northern | 172,518 (11.0) | 100,040 (10.4) |
| Zealand | 252,981 (16.2) | 159,016 (16.6) |
| Southern | 357,966 (22.9) | 216,872 (22.6) |
| **Vaccination priority groups** |  |  |
| Age priority | 1,155,301 (73.8) | 855,585 (89.3) |
| High risk of severe COVID-19 | 410,396 (26.2) | 103,048 (10.7) |
| **Comorbidities** |  |  |
| Chronic cardiac disorder | 304,829 (19.5) | 199,504 (20.8) |
| Malignancy | 165,323 (10.6) | 99,434 (10.4) |
| Autoimmune disorder | 160,074 (10.2) | 80,786 (8.4) |
| Diabetes | 110,655 (7.1) | 60,716 (6.3) |
| Psychiatric disorder | 113,087 (7.2) | 58,959 (6.2) |
| Chronic respiratory disorder | 64,276 (4.1) | 44,732 (4.7) |
| Asthma | 54,641 (3.5) | 28,797 (3.0) |
| Renal disorder | 26,322 (1.7) | 16,465 (1.7) |

| Characteristic | Entire study cohort<sup>b</sup> | Vaccinated with LP.8.1-containing vaccine |
| --- | --- | --- |
| Epilepsy | 15,477 (1.0) | 9,526 (1.0) |

<sup>a</sup> Entire study cohort consisted of all individuals aged 65 years and older or those at high risk of severe COVID-19 living in Denmark who had received previous COVID-19 vaccine doses. The study period was July 1, 2025, to December 3, 2025.
<sup>b</sup> Values are numbers (percentages) unless otherwise stated.
<sup>c</sup> 24 (0.0%) coded as "Region = other" in the entire study cohort and 10 (0.0%) among LP.8.1-vaccinated.

**Figure:**
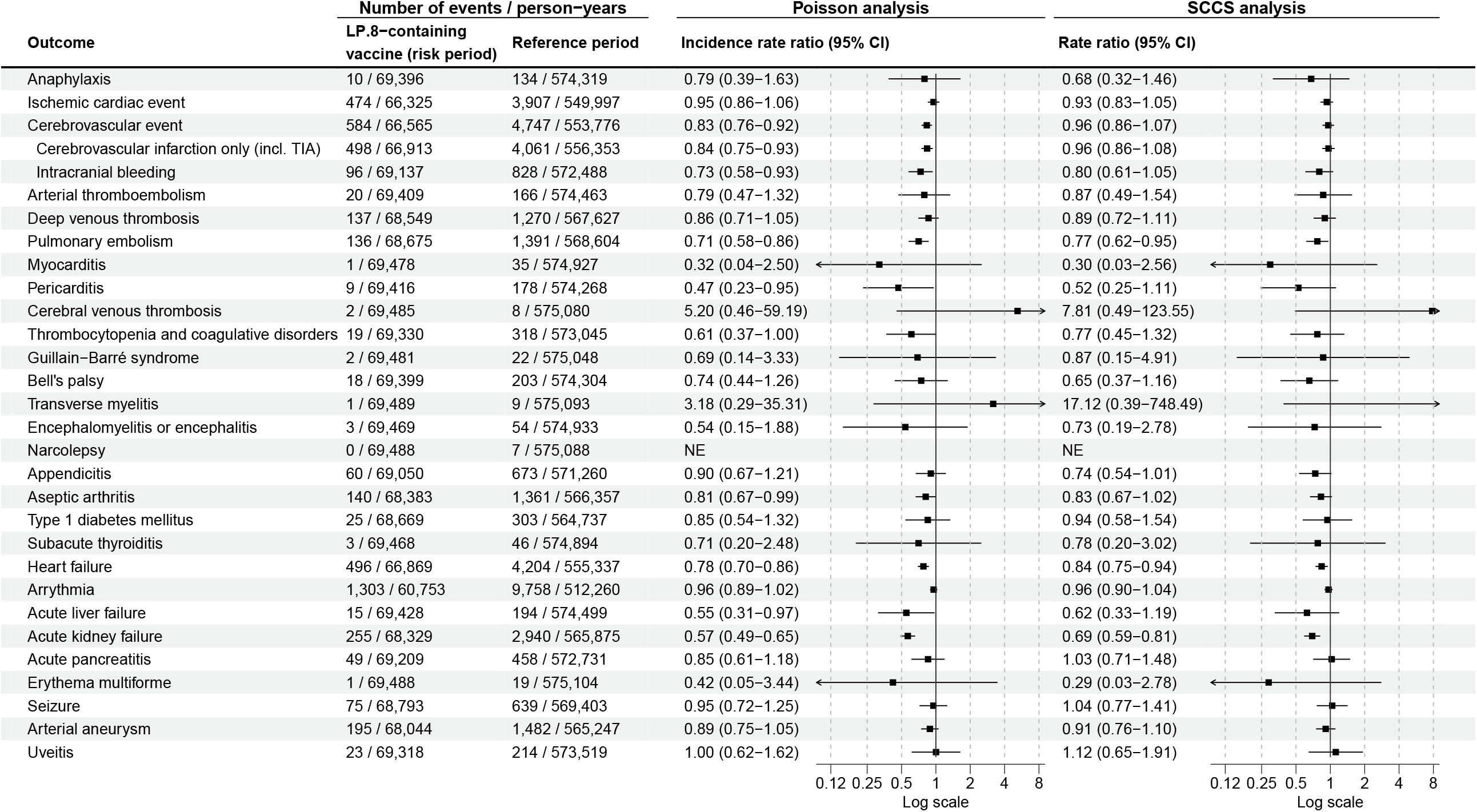
Risk of Adverse Events After Booster Vaccination With LP.8.1-Containing Vaccines The figure shows adjusted incidence rate ratios (IRRs) for 30 serious adverse events using Poisson regression and Self-Controlled Case Series (SCCS) analysis, respectively; rates observed during the 28-day risk period after vaccination with LP.8.1-containing SARS-CoV-2 mRNA vaccines are compared with rates during reference periods. Each outcome was studied separately, resulting in potential differences in denominators due to varying exclusions. NE, not estimable; TIA, transient ischemic attack.

## Discussion

In a nationwide cohort of more than 1.5 million adults, no increased risk of 30 adverse events was observed following vaccination with a LP.8.1-containing COVID-19 mRNA vaccine using two complementary analytical approaches.

Differences in ascertainment of adverse events between compared periods cannot be excluded, but in contrast to what was observed, would most likely bias toward increased risks. Including an SCCS analysis that inherently controls for time-invariant confounders reduces concern about residual confounding. Analyses were not adjusted for multiple testing, and some results suggested lower risk among vaccinated, indicating that a time-varying healthy vaccinee effect cannot be excluded.

### Data Sharing Statement

No additional data available. Owing to data privacy regulations in Denmark, the raw data cannot be shared. However, the data are available for research upon reasonable request to The Danish Health Data Authority and within the framework of the Danish data protection legislation and any required permission from Authorities.

## Author contributions

Dr Andersson and Ms Thiesson had full access to all the data in the study and takes responsibility for the integrity of the data and the accuracy of the data analyses.

*Concept and design*: All authors.

*Acquisition, analysis, or interpretation of data*: All authors.

*Drafting of the manuscript*: Hviid.

*Critical revision of the manuscript for important intellectual content*: All authors.

*Statistical analysis*: Andersson, Thiesson.

*Supervision*: Hviid.

