## Supplementary materials for "Adverse Events After LP.8.1-Containing COVID-19 mRNA Vaccines"

**eTable: Eligibility Criteria, and Outcome and Covariates Definitions**

**eFigure: Schematic Figure of the Study Design**

**eReferences**

**eTable: Eligibility Criteria, and Outcome and Covariate Definitions**

| **Variable** | **Details** |
| --- | --- |
| **Eligibility criteria** |  |
| Age of 65 years+ | The Civil Registration System.^1^ This core demography register holds the mandatory unique personal identifier for all permanent residents of Denmark allowing the cross-linkage of all national Danish registers. Age was defined as 2025 minus birthyear. |
| Vaccinated with previous COVID-19 vaccines | The Danish Vaccination Register.^2^ Defined as registered receipt of at least 3 previous COVID-19 vaccines (a primary series and a booster). In Denmark, the BNT162b2 and mRNA-1273 vaccines have been the primary COVID-19 vaccines with some AZD1222 use early on (AZD1222 use was stopped in March 2021). Use of other COVID-19 vaccines were rare and treated as censoring events. |
| High-risk group | The Danish Vaccination Register. Individuals 18+-years-of-age prioritized for vaccination due to being considered at high-risk of severe COVID-19. |
| **Outcomes** | **The National Patient Register.^3^ Both primary and secondary diagnoses recorded during any type of first-time hospital contact using ICD-10 codes.** |
| Anaphylaxis | ICD-10: T782, T783, T805, T886 |
| Ischemic cardiac event | ICD-10: I20-I251 |
| Cerebrovascular event | ICD-10: I60-66, G450-G453 |
| Cerebral infarction (incl. TIA) | ICD-10: I63, I64, G450-G453 |
| Intracranial bleeding | ICD-10: I60-62 |
| Arterial thromboembolism | ICD-10: I74 |
| Deep venous thrombosis | ICD-10: I80-82 (but not I800, I808C, or I821) |
| Pulmonary embolism | ICD-10: I26 |
| Myocarditis | ICD-10: I401, I408, I409, I418, I514 |
| Pericarditis | ICD-10: I300, I308, I309, I328 |
| Cerebral venous thrombosis | ICD-10: I636, I676 |
| Thrombocytopenia or coagulative disorders | ICD-10: D65, D683, D686, D688-689, D690, D693-D699 (but not D697 or D698A) |
| Guillain-Barré syndrome | ICD-10: G610 |
| Bell's palsy | ICD-10: G510 |
| Transverse myelitis | ICD-10: G373 |
| Encephalomyelitis or encephalitis | ICD-10: G040, G048, G049, G058, G361 |
| Narcolepsy | ICD-10: G474 |
| Appendicitis | ICD-10: K35-K37 |
| Aseptic arthritis | ICD-10: M10, M119, M130, M131, M139 |
| Type 1 diabetes mellitus | ICD-10: E10 |
| Subacute thyroiditis | ICD-10: E061 |
| Heart failure | ICD-10: I110, I420, I426-I429, I50, J81 |
| Arrhythmia | ICD-10: I44-I49 |
| Acute liver failure | ICD-10: K71, K72 |
| Acute kidney failure | ICD-10: D593, I12, I13, N00-N02, N04-N05, N08, N10, N141, N142, N144, N17, N19, R34 |
| Acute pancreatitis | ICD-10: K850, K853, K858, K859 |
| Erythema multiforme | ICD-10: L51 |
| Seizure | ICD-10: G40, G41 |
| Arterial aneurysm | ICD-10: I71, I72 |
| Uveitis | ICD-10: H20, H30 |
| **Covariates** |  |
| Sex | The Civil Registration System. Defined by registered sex (male or female). |
| Age | The Civil Registration System. Age was defined by year of study minus birthyear and categorized (18-39, 40-64, 65+). |
| Calendar time | The Civil Registration System. Treated as a time-varying covariate in monthly categories. |
| Region of residency | The Civil Registration System. Defined by the last registered address and categorized according to: Northern Denmark Region, Central Denmark Region, Region of Southern Denmark, Capital Region of Denmark, and Region Zealand. |
| Vaccination priority groups | The Danish Vaccination Register. The register holds information on governmentally prioritized COVID-19 vaccine groups assigned according to whether an individual was considered at being at high risk of severe COVID-19. |
| Comorbidities | The National Patient Register. The register holds information on all hospital contacts in Denmark. We defined comorbidity status as any registered primary or secondary diagnosis regardless of the hospital contact type between January 1, 2018 and study start. The number of comorbidities present in this pre-baseline period was categorized as 0, 1, or ≥2 comorbidities. |

*ICD-10 denotes International Classification of Diseases System, version 10; TIA, transient ischemic attack.*

**eFigure. Schematic Figure of the Study Design**

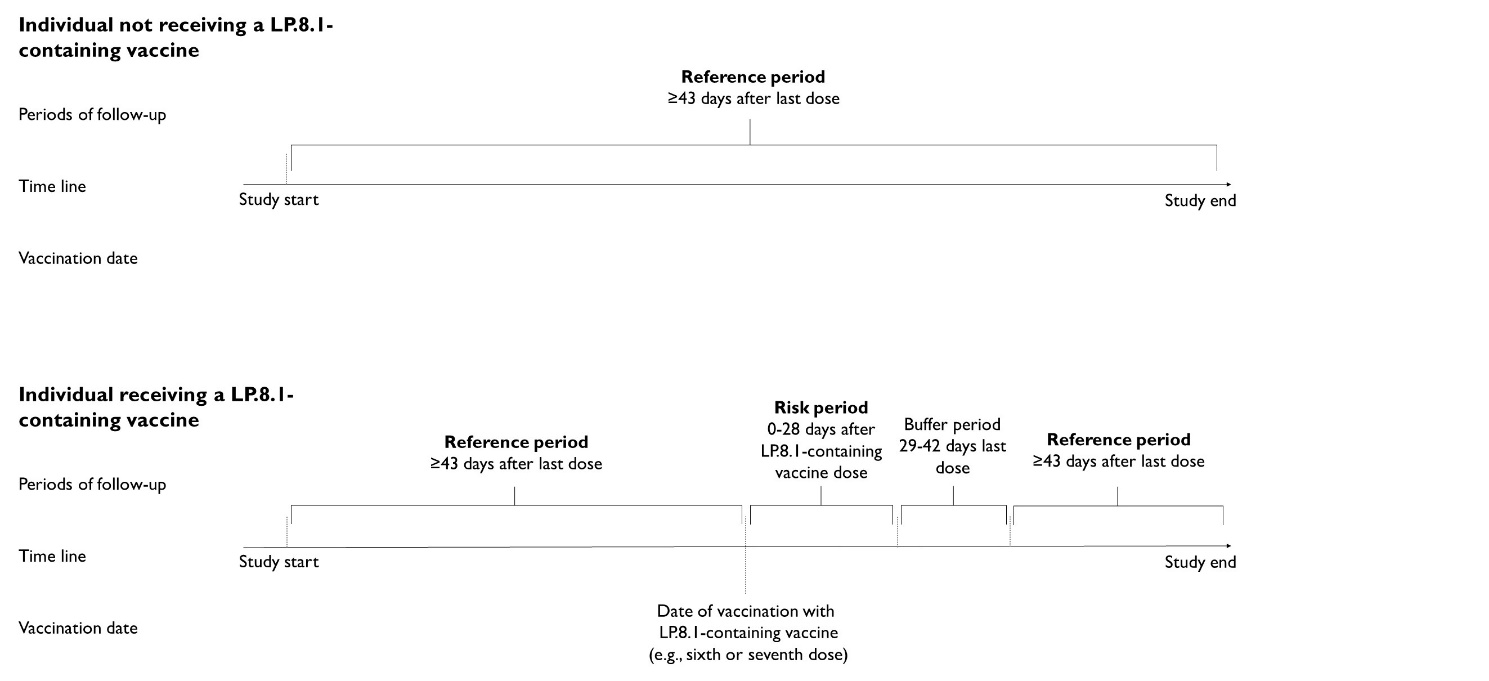

The risk period was from day 0 to day 28 following vaccination with an LP.8.1-containing vaccine. The reference periods consisted of ≥43 days following a vaccine dose from a previous season (up until the day before a potential LP.8.1-containing vaccination or study end) and ≥43 days following LP.8.1-containing vaccination (up until study end). Thus, individuals eligible for the study cohort but not receiving the LP.8.1-containing vaccine during the study period, contributed solely with reference period follow-up (in the ≥43 days following a vaccine dose from a previous season period).

Individuals could contribute with person-time during both the 28-day risk period and the reference periods. The days 29-42 after LP.8.1-containing vaccination was considered a buffer period and not included in the risk period or in the reference period follow-up.
